# Wastewater-based surveillance can be used to model COVID-19-associated workforce absenteeism

**DOI:** 10.1101/2023.01.22.23284878

**Authors:** Nicole Acosta, Xiaotian Dai, Maria A. Bautista, Barbara J. Waddell, Jangwoo Lee, Kristine Du, Janine McCalder, Puja Pradhan, Chloe Papparis, Xuewen Lu, Thierry Chekouo, Alexander Krusina, Danielle Southern, Tyler Williamson, Rhonda G. Clark, Raymond A. Patterson, Paul Westlund, Jon Meddings, Norma Ruecker, Christopher Lammiman, Coby Duerr, Gopal Achari, Steve E. Hrudey, Bonita E. Lee, Xiaoli Pang, Kevin Frankowsk, Casey R.J. Hubert, Michael D. Parkins

## Abstract

Wastewater-based surveillance (WBS) is a powerful tool for understanding community COVID-19 disease burden and informing public health policy. The potential of WBS for understanding COVID-19’s impact in non-healthcare settings has not been explored to the same degree. Here we examined how SARS-CoV-2 measured from municipal wastewater treatment plants (WWTPs) correlates with local workforce absenteeism. SARS-CoV-2 RNA N1 and N2 were quantified three times per week by RT-qPCR in samples collected at three WWTPs servicing Calgary and surrounding areas, Canada (1.3 million residents) between June 2020 and March 2022. Wastewater trends were compared to workforce absenteeism using data from the largest employer in the city (>15,000 staff). Absences were classified as being COVID-19-related, COVID-19-confirmed, and unrelated to COVID-19. Poisson regression was performed to generate a prediction model for COVID-19 absenteeism based on wastewater data. SARS-CoV-2 RNA was detected in 95.5% (85/89) of weeks assessed. During this period 6592 COVID-19-related absences (1896 confirmed) and 4,524 unrelated absences COVID-19 cases were recorded. Employee absences significantly increased as wastewater signal increased through the pandemic’s waves. Strong correlations between COVID-19-confirmed absences and wastewater SARS-CoV-2 signals (N1 gene: r=0.824, p<0.0001 and N2 gene: r=0.826, p<0.0001) were observed. Linear regression with adjusted R^2^-value demonstrated a robust association (adjusted R^2^=0.783), when adjusted by 7 days, indicating wastewater provides a one-week leading signal. A generalized linear regression using a Poisson distribution was performed to predict COVID-19-confirmed absences out of the total number of absent employees using wastewater data as a leading indicator (P<0.0001). We also assessed the variation of predictions when the regression model was applied to new data, with the predicted values and corresponding confidence intervals closely tracking actual absenteeism data. Wastewater-based surveillance has the potential to be used by employers to anticipate workforce requirements and optimize human resource allocation in response to trackable respiratory illnesses like COVID-19.

**Highlights:**

- WBS is a useful strategy for monitoring infectious diseases in workers
- SARS-CoV-2 RNA in wastewater correlated with workforce absenteeism
- Workplace absenteeism secondary to COVID-19 can be predicted using WBS
- WBS can be used by employers to anticipate and mitigate work force absenteeism

**Graphical abstract:** 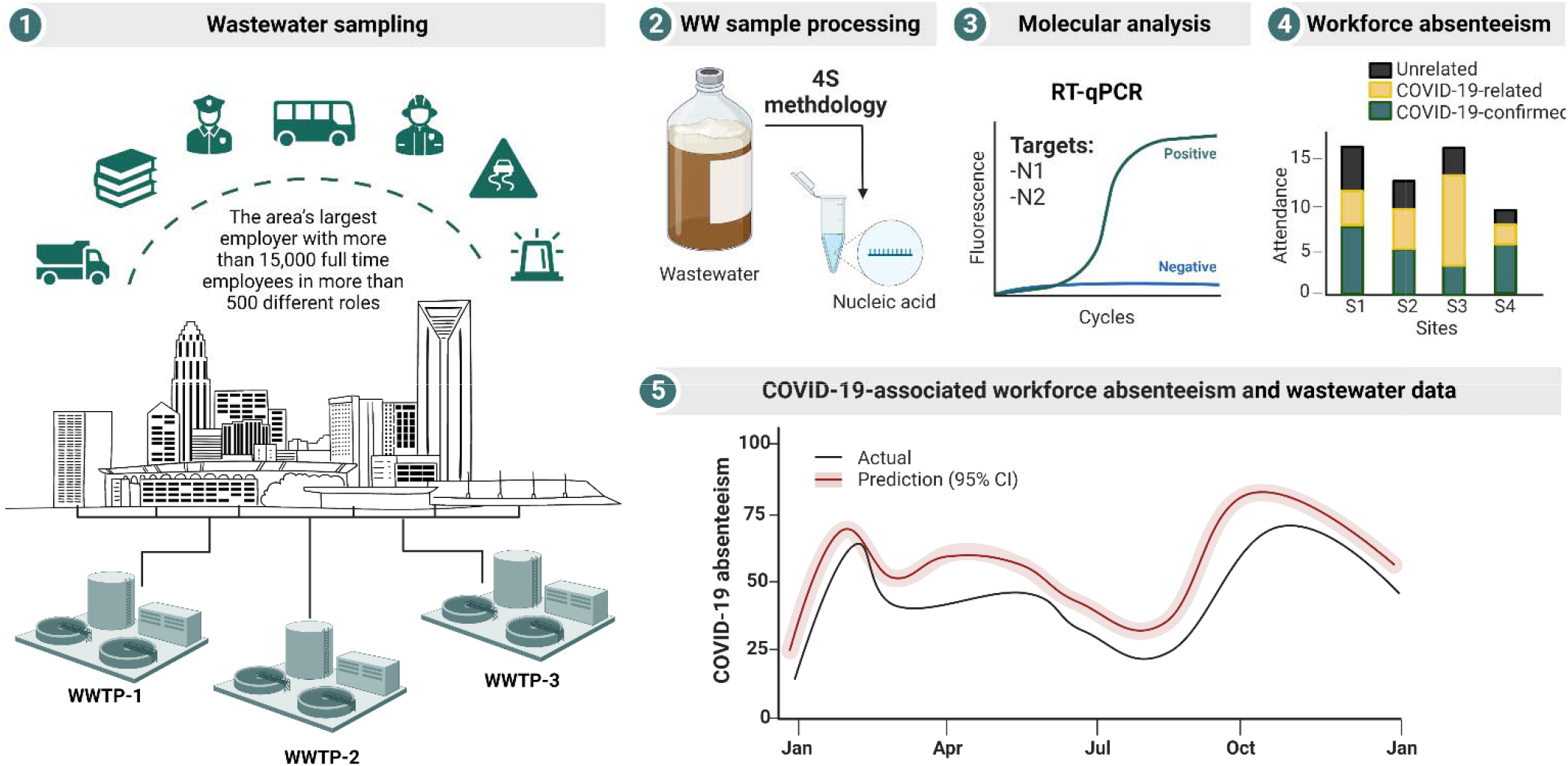

## 1. INTRODUCTION

Wastewater-based surveillance (WBS) has emerged as a useful strategy enabling real-time monitoring of the incidence and prevalence of several infectious diseases at the population level (Arora et al. 2022; Asghar et al. 2014; Hellmér et al. 2014). The strength of WBS relates to its ability to provide longitudinal objective and inclusive information on the burden of disease in monitored populations that is independent of clinical testing and other healthcare system resources. Furthermore, WBS represents an intrinsically comprehensive and inclusive strategy that captures all individuals served by the sewer system, irrespective of symptoms or capacity to seek testing (Tang et al. 2020; Wu et al. 2020; Amirian 2020). With the emergence of the Severe Acute Respiratory Syndrome Coronavirus 2 (SARS-CoV-2) pandemic and the recognition that its genomic viral RNA is shed in the feces of infected individuals, WBS to monitor for COVID-19-related disease burden became an intense research focus (Kitajima et al. 2020; Wölfel et al. 2020; Ling et al. 2020; Medema et al. 2020).

Researchers around the world have demonstrated WBS can provide a leading indicator of many clinically relevant parameters, including; community diagnosed COVID-19 cases, COVID-19-related hospitalizations and ICU admissions, and even deaths (Acosta et al. 2021; Acosta et al. 2022; Peccia et al. 2020; Randazzo et al. 2020; Miyani et al. 2021; Zhu et al. 2021; Xiao et al. 2021; Zhan et al. 2022; Galani et al. 2022; Hegazy et al. 2022). Recognizing the tremendous potential of WBS to help shape and inform public health policy and actions including healthcare resource allocation, WBS programs have developed all over the world (Naughton et al. 2021). However, the utility of WBS-derived data to other areas of society beyond public health has not been well explored. Given the profound influence workplace COVID-19 related absenteeism has on the ability of private and public organizations to fulfill their mandate and deliver services, we sought to explore its relationship with SARS-CoV-2 signals measured in municipal wastewater. Our data demonstrate that workplace absenteeism at the largest employer in Canada’s fourth largest city correlated strongly with and can be predicted using wastewater data.

## 2. MATERIAL AND METHODS

### 2.1 Wastewater collection and processing

Wastewater was collected from all three wastewater treatment plants (WWTPs) servicing Calgary and surrounding communities, Alberta, Canada. City of Calgary staff collected samples from WWTP-1, WWTP-2 and WWTP-3 between June 29, 2020, and March 28, 2022. Autosamplers were programed to collect 24-hour composite samples at each WWTP (Acosta et al. 2022), and delivered to the City of Calgary lab where a sub-sample was aliquoted from the bulk sample. The sub-samples were immediately transported to the University of Calgary’s Advancing Canadian Water Assets (ACWA) laboratory for processing and nucleic acid extraction as detailed elsewhere (Whitney et al. 2021; Acosta et al. 2021). All samples were spiked with an external bovine coronavirus control (BCoV; Bovilis, Merck) to ensure consistency in processing. Extracted nucleic acids were transported to the Health Sciences Center at the University of Calgary on dry ice. This project was approved by the Conjoint Regional Health Ethics Board (REB20-1252).

RT-qPCR was used to quantify the SARS-CoV2 RNA in wastewater using a QuantStudio-5 Real-Time PCR System (Applied Biosystems). SARS-CoV-2 RNA concentration was measured utilizing nucleocapsid N1 and N2 targets (CDC 2020b; Acosta et al. 2021), with both genes being analyzed for comparison. 2019-nCoV_N positive control plasmid (Integrated DNA Technologies, Catalogue # 10006625) was used as an RT-PCR standard. The BCoV external control was quantified following a previously described protocol (Acosta et al. 2021). Each sample was measured in triplicate, and each RT-qPCR run included no-template controls (NTCs). Samples were considered positive for the presence of SARS-CoV-2 RNA-target if amplification passed a detection cycle threshold in <40 cycles (Acosta et al. 2021; Randazzo et al. 2020). Gene abundances for both targets (i.e., N1 and N2) per volume of wastewater for each WWTP were calculated and then combined as a single city-wide metric encompassing all three WWTPs using a validated approach that combines flow rate at each WWTP (Acosta et al. 2022). The flow rate of each WWTP was used to normalize the SARS-CoV-2 RNA concentration and reported as viral gene abundance per day.

### 2.2 Determining absenteeism rates among municipal employees

Calgary is Canada’s fourth largest city (1.3 million residents) and third most ethnically diverse. The City of Calgary is Calgary’s largest individual employer, with more than 15,000 full-time employees in more than 500 different roles (i.e., Calgary Transit >3000 staff, Calgary Police Services >2600 staff, etc.)(City-of-Calgary 2022). During the pandemic, data on employee absenteeism was monitored by the Calgary Emergency Management Agency (CEMA) team and was classified as COVID-19-related, COVID-19-confirmed or unrelated to COVID-19. Unrelated cases were any cause for absenteeism that was not directly related to COVID-19 (i.e., injuries, etc.) (Figure 1 and Table 1S). Cases that were COVID-19-related included those manifesting from potential exposure to a suspected/confirmed individual at home or work and those employees with respiratory symptoms themselves, whether confirmed to be COVID-19 or not, thereby excluding the employee from safely returning to the workplace. COVID-19-confirmed were the subset of COVID-19-related cases that were ultimately verified with a clinical diagnostic test. Clinical case confirmation came from the single provincial health provider Alberta Health Services, using standard clinical testing protocols evaluating RT-qPCR of nasopharyngeal/nasal swabs (Feb 2020-March 2022) and then supplemented with self-collected home diagnostic tests (Dec 2021-March 2022). Anonymized weekly numbers of absent employees due to COVID-19-related were collected over 89 weeks (June 2020 to March 2022).

**Figure 1.**
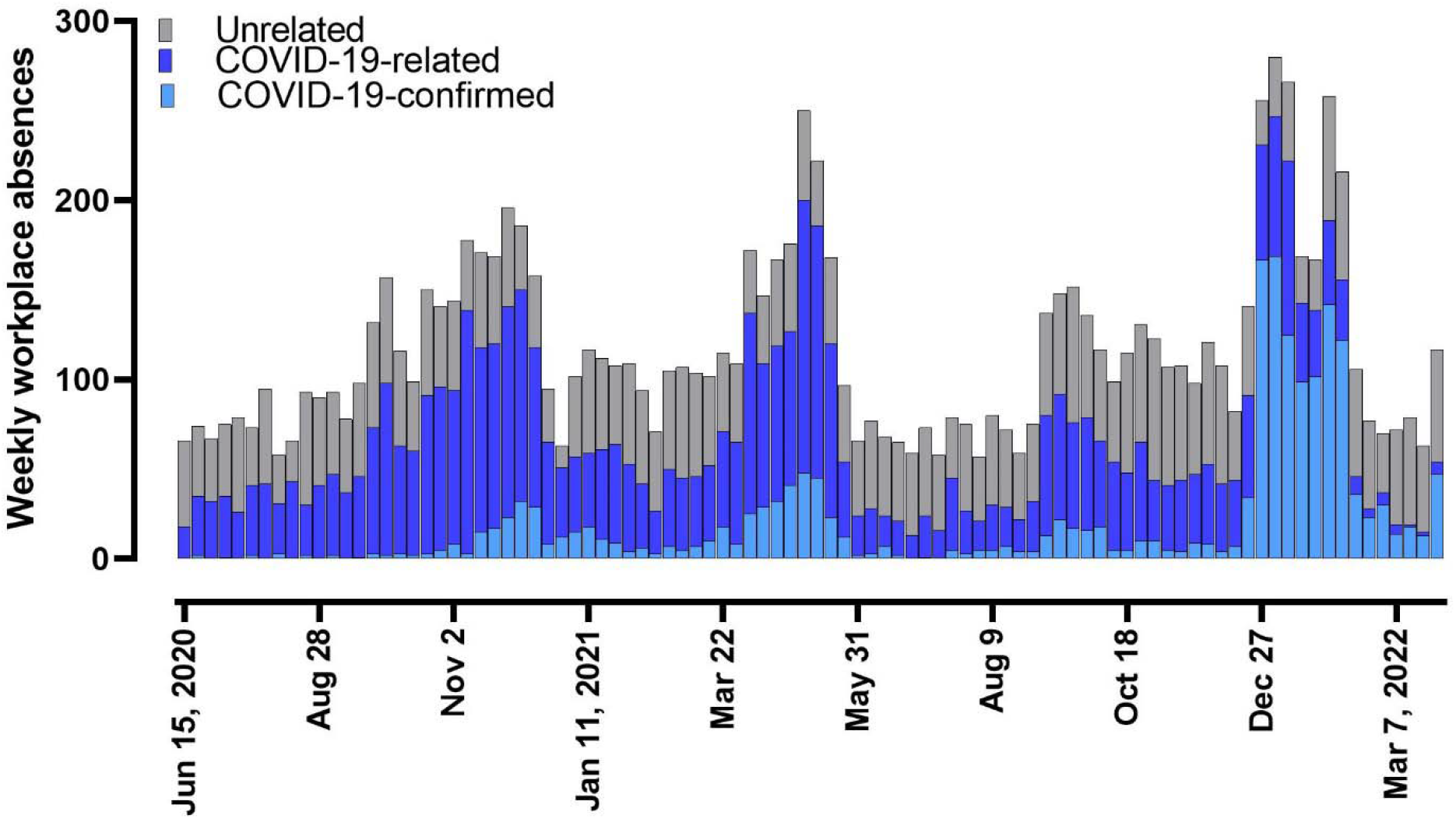
Municipal workforce absenteeism from June 2020 to March 2022. Employee absences as measured weekly categorized as unrelated (grey bars), or COVID-19-related (blue bars). COVID-19-related absences (dark blue bars) include those associated with known exposure to an infected individual or symptoms. COVID-19-confirmed absences (light blue bars) correspond to a clinical PCR test result.

Waves of COVID-19 in Calgary were defined by Alberta Health (AH) and AHS Public Health surveillance teams based on active case counts. During the 89-week study period, six COVID-19 waves were reported in Alberta: wave-1 (March 8, 2020, to July 10, 2020), wave-2 (July 10, 2020, to February 25, 2021), wave-3 (February 25, 2021, to July 1, 2021), wave-4 (July 1, 2021, to December 12, 2021), wave-5 (December 12, 2021, to March 6, 2022) and wave-6 (March 6, 2022, to June 26, 2022).

### 2.3 Statistical analysis

All data analyses were performed in GraphPad Prism-8 software (La Jolla, CA) and R (V4.0.4) using the olsrr-package. The 7-day rolling average of the viral gene abundance per day was established using aggregate city-wide SARS-CoV-2 burden in wastewater determined three times each week at each WWTPs for both N1 and N2 gene targets used for quantifying SARS-CoV-2 RNA. Spearman’s rank correlation was used to evaluate the relationship between the weekly averaged viral gene abundance per day in wastewater and absenteeism attributed to confirmed COVID-19 among municipal employees. Cross-correlation function (CCF) analysis was performed using SARS-CoV-2 RNA concentration (gene abundance/day) values as the independent variable and COVID-19-confirmed absenteeism as the dependent variable. Linear regression with R^2^ was performed to evaluate how much the measured wastewater signal could explain the variation of COVID-19-confirmed absences. Generalized Poisson regression model was performed to predict the number of COVID-19-confirmed absences out of the total number of absent employees using the wastewater signal using 1 lag (week) and the number of absent employees. One week lag was chosen because COVID-19-confirmed absences had the strongest positive correlation with SARS-CoV2 RNA concentration in wastewater collected 1 week prior – and is the most clinically relevant. The Poisson regression model can be formulated as;

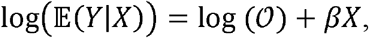

where *Y* is the number of cases of COVID-19-confirmed absences out of the total number of absent employees in the current week, *X* is the wastewater signal (e.g., N1 or N2) 1 week prior, 𝒪 is the offset term and represents the number of absent employees in the current week, and 𝔼(*Y*|*X*) is the expected COVID-19 absenteeism given wastewater signals. It can be assumed *Y* follows a Poisson distribution because absenteeism data is a non-negative integer. When performing predictions, standard errors of predictions can be calculated using the *predict*.*glm* function in R (R Core Team, 2013). We then obtain the 95% confidence interval of one predicted value 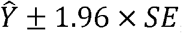, where SE is the standard error of the predicted value. The predictive model was built on 2020 data.

## 3. RESULTS

### 3.1 COVID-19 absenteeism among municipal staff during the pandemic

Between June 15, 2020 and March 28, 2022 there were 11,116 municipal staff absences. Of those, 4,524 were unrelated to COVID-19, with the other 6,592 absences (59.3%) classified as COVID-19-related (Figure 1). Among COVID-19-related absences, 1,896 were confirmed by clinical testing, for a 20.2% weekly average positivity rate. There was an uneven distribution of COVID-19-confirmed absences during the different pandemic waves: Wave-1, 3/120 (2.5 %); wave-2, 248/2274 (10.9 %); wave-3: 317/1441 (21.9%); wave-4: 180/1077 (16.7%); wave-5: 1056/1573 (67.13%) and wave-6: 92/107 (85.9%). The maximum number of weekly COVID-19-confirmed absences (i.e., 169) was observed during the first week of 2022, which corresponded to wave-5 (Omicron/BA.1) in Calgary (Figure 1).

### 3.2 Wastewater measured SARS-CoV-2 across the City

Wastewater samples were analyzed up to three times per week from June 29, 2020, to March 28, 2022 (Supplementary Figure 1). A total of 524 wastewater samples were collected at Calgary’s wastewater treatment plants (WWTPs) (WWTP-1: 174, WWTP-2: 176, and WWTP-3: 174). Of the total wastewater samples tested using the N1, and N2 assays, 517 (98.7%) samples were positive for N1 and 496 (94.7%) were positive for N2. We combined wastewater-measured SARS-CoV-2 RNA concentration into a single city-wide metric after the daily flow rate of each WWTP was used to normalize the RNA concentration and reported as viral gene abundance per day. The SARS-CoV-2 RNA combined signal measured with either N1 or N2 targets was used for all analyses (Figures 2 and 3). SARS-CoV-2 RNA concentration was low (black lines) during the summer to fall of 2020 (June to November 2020) and rapidly increased during Alberta’s second wave in December 2020.

**Figure 2.**
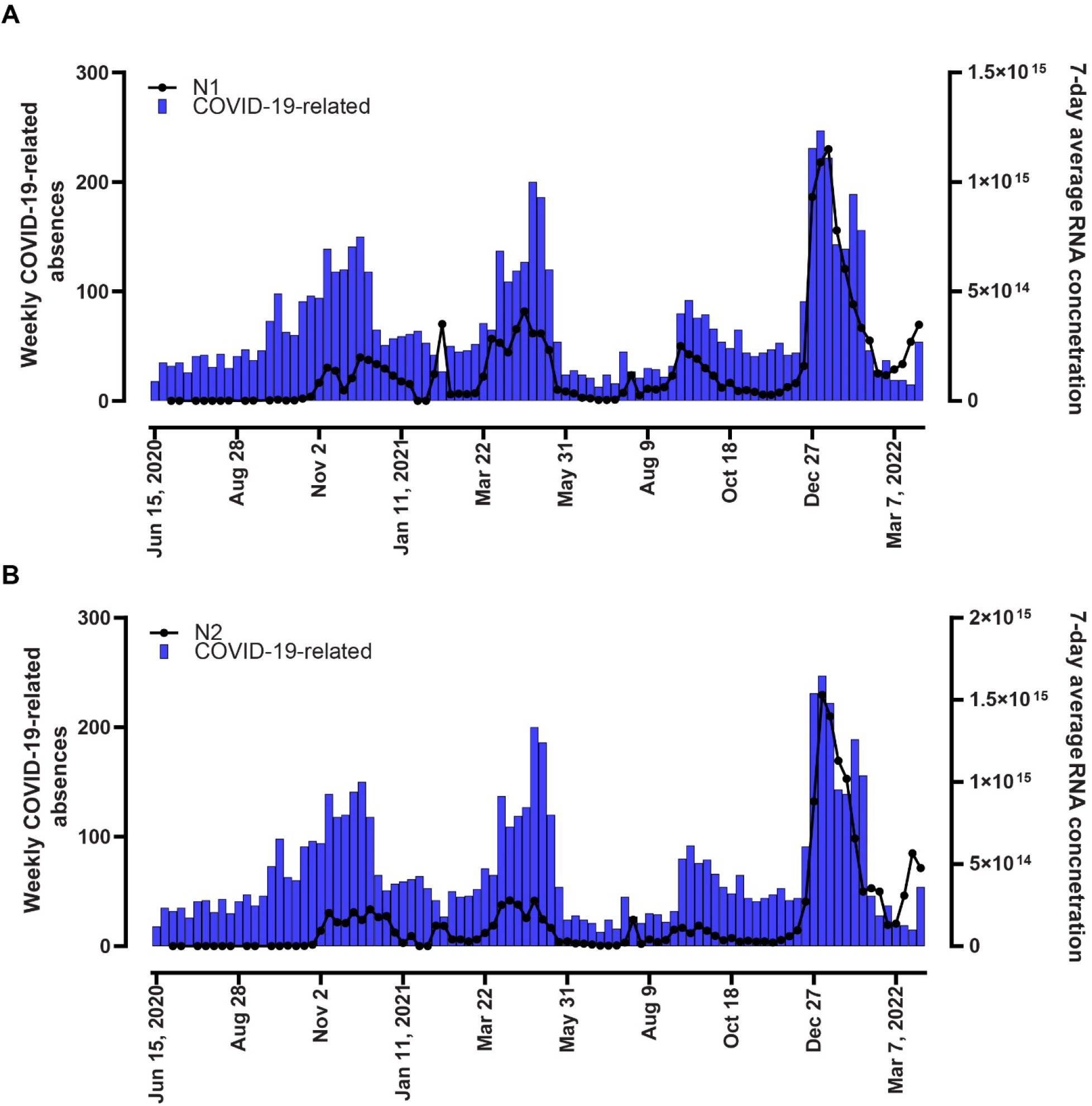
Comparing SARS-CoV-2 RNA concentration in wastewater to <u>COVID-19-related</u> municipal absenteeism. The figure demonstrates the correlation between the weekly wastewater averaged gene abundance per day and the weekly rate of employee absenteeism attributed as COVID-19-related (dark blue bars). Flow-adjusted values for wastewater concentrations of SARS-CoV-2 genes N1 (A) and N2 (B) over the study duration are shown as black doted lines.

**Figure 3.**
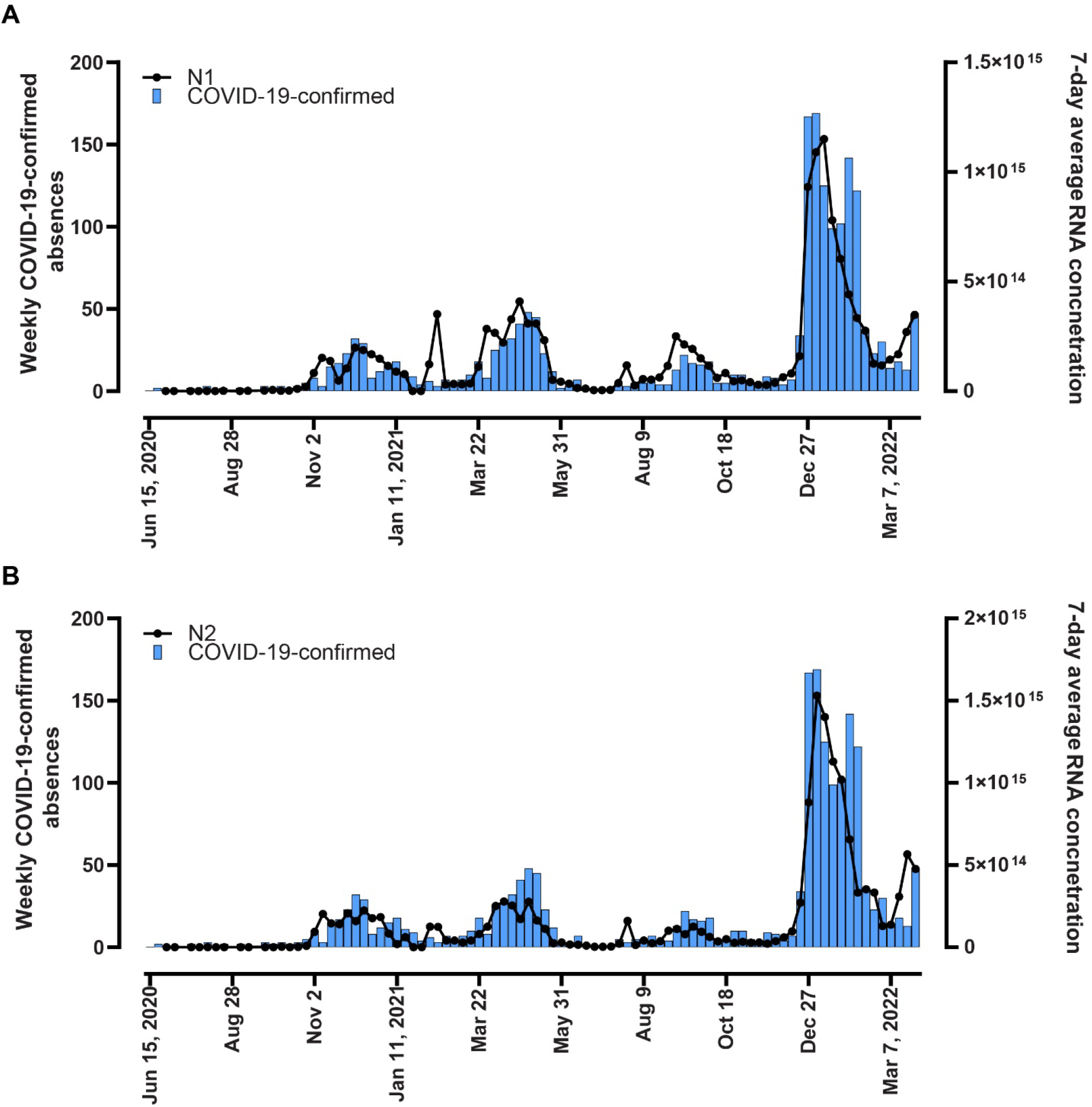
Comparison of SARS-CoV-2 RNA concentration in wastewater to municipal absenteeism rates attributable to <u>confirmed COVID-19</u>. The figure demonstrates the correlation between the weekly wastewater averaged gene abundance per day and the number of new municipal employee absenteeism rates attributed to <u>confirmed COVID-19</u> (light blue bars). Flow-adjusted values for wastewater concentrations of SARS-CoV-2 genes N1 (A) and N2 (B) over the study duration are shown as black doted lines.

### 3.3 Correlating absenteeism among municipal service workers and wastewater measured SARS-CoV-2 RNA

The weekly number of COVID-19-related (Figure 2) and COVID-19-confirmed absences (Figure 3) declined from mid-January to March 2021, accompanied by a decrease in the total SARS-CoV-2 wastewater RNA concentration. The changing dynamics of employee absenteeism during waves 3 to 6 were mirrored by the changes in SARS-CoV-2 wastewater RNA signal. When weekly averaged N1, and N2 SARS-CoV-2 RNA concentration in wastewater were compared with the total workforce absences (i.e., COVID-19-related, and unrelated), and COVID-19-confirmed absences, we found a relationship between employee absenteeism and city SARS-CoV-2 wastewater levels. Because city-wide wastewater levels were collected up to three times per week (on Mondays, Wednesdays and Thursdays), weekly averaged signals were established and paired with the weekly number of total absences. A moderate association between total absences and the SARS-CoV-2 genomic RNA signal in wastewater (N1: Spearman correlation *r*=0.529, 95% CI for correlation coefficient: 0.355 - 0.667, P<0.0001; N2: Spearman correlation *r*=0.495, 95% CI for correlation coefficient: 0.314 - 0.641,) was found. When we assessed the association between weekly COVID-19-related absence and wastewater measured N1 or N2 signal, we observed a similar trend [(N1; Spearman correlation *r*=0.518, 95% CI for correlation coefficient: 0.343 - 0.660, P<0.0001) (Figure 2A) and N2 (Spearman correlation *r*=0.479, 95% CI for correlation coefficient: 0.295 - 0.629, P<0.0001)] (Figure 2B) signal. Notably, N1 and N2 measured SARS-CoV-2 explained a smaller proportion of COVID-19-related absenteeism at the beginning of the pandemic (waves 1 and 2) relative to later in the monitoring period (waves 3 to 6). To evaluate the effect of the COVID-19 pandemic waves over time, the weekly SARS-CoV-2 RNA concentration was paired with weekly COVID-19-related absenteeism as a function of the waves, and the cross-correlation function was applied. Wave-1 and 6 were not included in the analysis because too few data points were available. The correlation between absenteeism and SARS-CoV-2 concentration was the strongest when using - 1 week lag (correlation value at -1 lag for wave-2: 0.516, wave-3: 0.869, wave-4: 0.74; and wave-5: 0.494) (Supplementary Figure 2). The number of spikes was smaller in later waves, which could be explained by the fact that as the pandemic evolved there could be a higher rate of vaccination and immunity among the employees.

When Spearman’s rank correlation was performed to assess COVID-19-confirmed absenteeism as a function of wastewater viral RNA levels, even stronger associations were observed (Figure 3). There was a strong correlation between weekly COVID-19-confirmed absenteeism and the weekly average SARS-CoV-2 signal for both N1 (Spearman correlation *r*=0.824, 95% CI for correlation coefficient: 0.741 - 0.883, P<0.0001) (Figure 3A) and N2 (Spearman correlation *r*=0.826, 95% CI for correlation coefficient: 0.744 - 0.884, P<0.0001) (Figure 3B). Correlation coefficients showed that SARS-CoV-2 RNA concentration in wastewater increased in accordance with employee absenteeism rates. Overall, there was a linear association between WBS data and COVID-19-confirmed absenteeism. To evaluate the temporal effect, the weekly SARS-CoV-2 RNA concentration was paired with weekly COVID-19-confirmed absences, and the cross-correlation function applied. We found a positive correlation between the SARS-CoV-2 concentrations, measured with either N1 (correlation value at -1:0.800, 0: 0.900 and 1:0.815 lag) or N2 (correlation value at -1: 0.822, 0: 0.888 and 1: 0.843 lag) targets and absenteeism within a difference of − 1 and 1 lag (weeks) (Supplementary Figure 3).

### 3.4 Predictive model for COVID-19-confirmed absenteeism

Given the strongest correlation between COVID-19-confirmed absenteeism and wastewater measured SARS-CoV-2 N1 and N2 (Figure 3), we proceeded with modelling using this parameter alone. A linear regression model with adjusted R^2^ was performed to investigate the goodness of fit of a predictive model using a normal distribution. The model had a very strong predictive capacity using either SARS-CoV-2 gene target in wastewater: N1 (adjusted R^2^ = 0.783 (for linear regression model); P-value < 0.0001) and N2 (adjusted R^2^ = 0.756 (for linear regression model); P-value < 0.0001). Nearly 80% of the total variation in COVID-19-confirmed absenteeism is explained by the wastewater signals measured by either target. Autocorrelation, partial autocorrelation and normality (Quantile-quantile) plots of the residuals of the model suggested that the correlation among residuals is reasonable small and there was not obvious outlier in the data (Supplementary Figure 4). A generalized linear regression with a Poisson distribution was performed to better understand what proportion of COVID-19-confirmed absenteeism could be estimated by the wastewater signal. As the N1 wastewater signal gave a better Adjusted R^2^ coefficient and N1 and N2 correlated well (Supplementary Figure 5), we proceeded only with this prediction model measuring the regression of COVID-confirmed absenteeism (Y) on SARS-CoV-2 N1 measured in wastewater collected one week prior (1-week leading signal). The predictive model was built on 2020 data, and then it was used to predict COVID-19-confirmed absenteeism with associated 95% confidence intervals from January 2021 to March 2022 (Figure 3A). We found that the model can predict COVID-confirmed absenteeism using SARS-CoV-2 N1 abundance in wastewater. The model predictions closely track the actual absences (Figure 4). As shown in Figure 4, the interval width is relatively small compared to the scale of predicted Y values, which suggests the predictions and estimated regression coefficients have low variations and the predictions are reliable. The final model predictions track the overall trend of actual COVID-19 absences very well.

**Figure 4.**
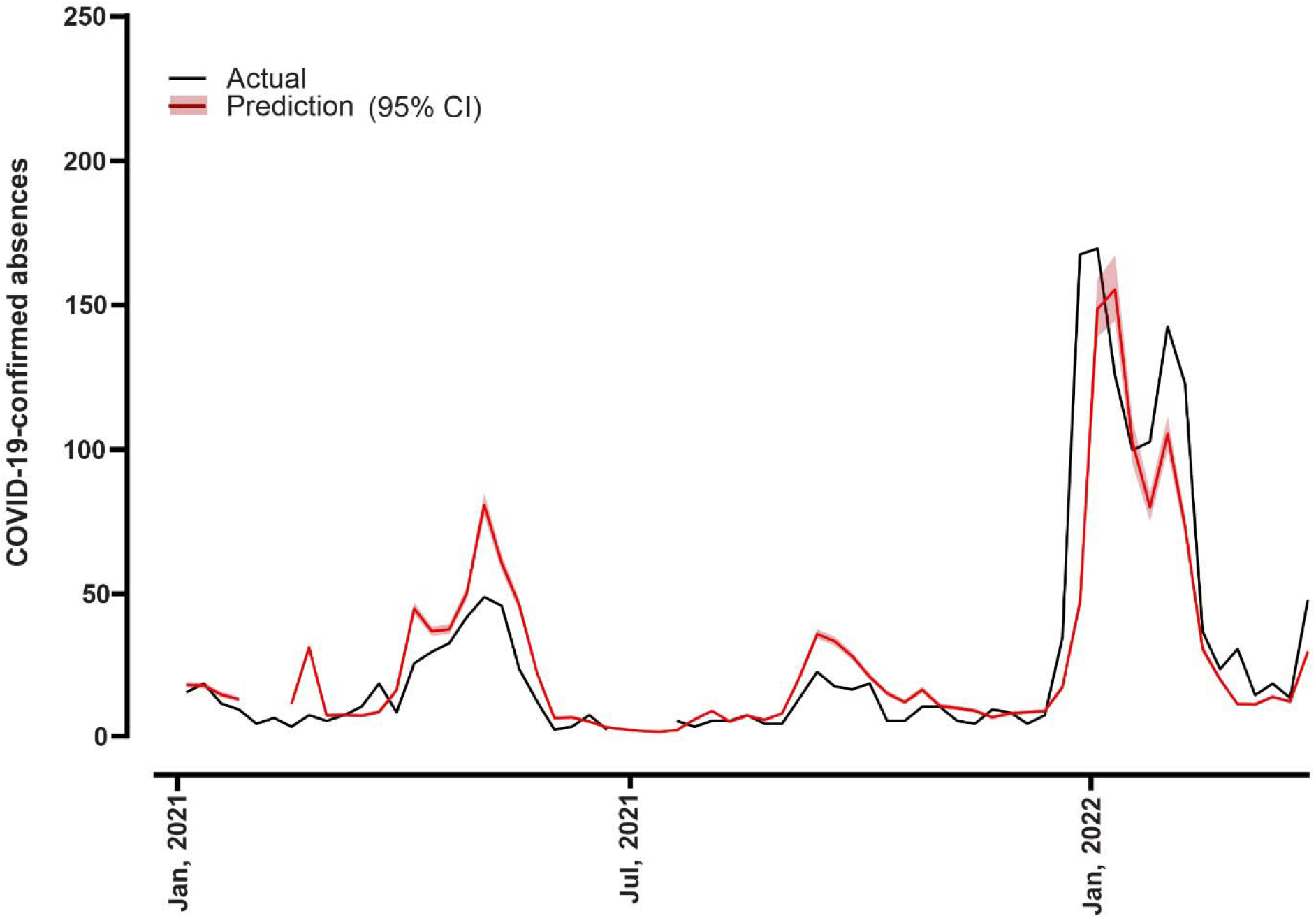
Predictive model for COVID-19 absenteeism in the municipal workforce. Plot of Poisson regression model (95% confidence limits) for the prediction of COVID-19 absenteeism amongst municipal workers as determined through city-wide SARS-CoV-2 N1 gene abundance measured in the preceding week’s wastewater. The model was built using the 2020 data and verified using COVID-19-confirmed absenteeism data from January 2021 to March 2022

## 4. DISCUSSION

To date, WBS studies have primarily focused on understanding the temporal changes in COVID-19 in communities by measuring SARS-CoV-2 RNA concentration in wastewater derived from WWTP (Pang et al. 2022; Acosta et al. 2022; Ahmed et al. 2020; de Sousa et al. 2022; Hasan et al. 2021; Karthikeyan et al. 2021; Wartell et al. 2022; Tandukar et al. 2022; Acosta et al. 2021; Gonçalves et al. 2021; Wang et al. 2020; Zhou et al. 2021; Gibas et al. 2021; Betancourt et al. 2021; Dai et al. 2022). These data have enabled evidence-informed decisions by public health officials as well as members of the public. Few studies, however, have looked to harness wastewater-based surveillance as a tool to influence business policy. There have been a modest number of reports of wastewater from select high-risk facilities such as hospitals and university dormitories being monitored for SARS-CoV-2 RNA enabling decisions on a more local level (Tandukar et al. 2022; Acosta et al. 2021; Gonçalves et al. 2021; Wang et al. 2020; Zhou et al. 2021; Gibas et al. 2021; Betancourt et al. 2021; Dai et al. 2022). However, studies assessing how WBS may be harnessed to affect other societal factors remain lacking. Absenteeism is one such area relevant to all industries and employers. For example, the high and unpredictable rates of workforce absenteeism associated with the COVID-19 pandemic have stifled job growth and economic recovery (Faramarzi et al. 2021).

COVID-19 absenteeism has predominantly been explored based on its impact on the healthcare workforce. Studies have invariably demonstrated a marked increase in absenteeism rates relative to pre-pandemic periods (de Paiva, Dos Santos, and de Lima Dalmolin 2022; Grandi, Silva, and Barbosa 2022; Challener et al. 2021). Indeed, these have been significant contributing factors manifesting in record-high rates of fatigue, burnout and workers prematurely leaving the workforce relative to pre-pandemic times (Maunder et al. 2021; Murthy 2022). However, absenteeism in general populations have also had profound socioeconomic impacts. Some of the best data tracking these trends come from the US Centres for Disease Control’s National Institute for Occupational Safety and Health (NIOSH), which monitors the prevalence of health-related absenteeism among full-time workers. Higher absenteeism rates early in the pandemic relative to prior flu seasons were confirmed (peaking in December 2021-January 2022 at 5.4% vs ∼2.5%) (CDC 2020a; Groenewold et al. 2020). Absenteeism was most significantly increased in production, service provision and transportation industries. Critically, several categories of “essential workers” (those whose jobs were considered critical to societal functioning and cannot be performed remotely) (Government-of-Canada 2021) include those working for municipalities (i.e., transportation, social services and policing) and have been disproportionally impacted with COVID-19 based on serosurveys (Shah et al. 2022).

Here, we found a positive relationship between weekly SARS-CoV-2 RNA concentrations in wastewater treatment plants servicing Calgary and confirmed absences reported in the week following sampling collection, confirming that wastewater provides a leading signal for workplace absenteeism. This relationship was not as strong at the beginning of the pandemic (waves 1 and 2) compared to later waves. This may be in part relate to the lower incidence of COVID-19 and greater uncertainty at the beginning of the pandemic as wastewater-based surveillance has been shown to be sensitive to an incidence rate of ∼2/100,000 populations (Li et al. 2023; Acosta et al. 2022). In addition, the detection of SARS-CoV-2 RNA in wastewater using either gene N1 or N2 showed a good correlation. Still, as the N1-signal gave a better Adjusted R^2^ coefficient, it was only included in the final prediction model. Previous studies utilizing wastewater data had focused on models to describe, estimate, and interpret SARS-CoV-2 RNA concentrations across general populations (de Sousa et al. 2022; Karthikeyan et al. 2021; Krivoňáková et al. 2021; Vallejo et al. 2022; Ahmed et al. 2020; Hasan et al. 2021; Saththasivam et al. 2021). Simulation of infection prevalence based on the viral RNA concentration in sewage has been developed using different modeling approaches, including Monte Carlo simulations, susceptible-exposed-infectious-recovered (SEIR) model, autoregressive integrated moving average (ARIMA) models (de Sousa et al. 2022; McMahan et al. 2021; Karthikeyan et al. 2021; Wu et al. 2022). Here, the estimation of the number of absent workers due to COVID-19 infections was performed via a Poisson regression model. Other studies also have shown the trends and strong correlation of clinical data with the SARS-CoV-2 RNA abundance in wastewater on a weekly basis (Wartell et al. 2022; Fahrenfeld et al. 2022; Halwatura et al. 2022; Krivoňáková et al. 2021).

Strategies to mitigate the impact of absenteeism in the workforce generally involve worker replacement, redistributing work among healthy workers (extra shifts and overtime), and new recruitment and training (Aguilar et al. 2021). However, tools that independently forecast future absenteeism rate changes based on objective and independent data, such as that delivered by WBS still need to be improved. In addition to ensuring workplaces have the human resources to fulfill their mandate, such a model can be used to optimize costs associated with a dynamic workforce. Indeed, the indirect costs related to absenteeism along with the workforce during the COVID-19 pandemic were estimated by the Integrated Benefits Institute to have cost US employers almost $1 billion each week (paid as a combination of excess overtime and other employee benefits, or sick leave and disability) (Schiavo 2021). Another important aspect related to absenteeism at the workplace is its impact on shift scheduling. Alongside wages, shift scheduling unpredictability may have negative impact on health and economic insecurity from the employee perspective (Harknett, Schneider, and Irwin 2021). Therefore, models that establish thresholds in wastewater SARS-CoV-2 signal one week in advance could be used as triggers for internal actions like absence recovery policies (i.e., holdover overtime, call-ins, cross-training, and temporary workers) (Easton and Goodale 2005; Maass et al. 2017; Easton 2011; Harknett, Schneider, and Irwin 2021). The prediction models, such as the one we developed, could be used to identify wastewater SARS-CoV-2 thresholds that can be used to trigger internal compensatory actions.

This study has several notable limitations. First, the estimation of the true rate of COVID-19 absenteeism in our study population could be affected by different factors. Asymptomatic infection in some workers or clinical testing hesitancy could result in wastewater signals without corresponding absences from work. Second, owing to changes in testing availability and the perception of COVID-19-associated risk among populations, the proportion of positive COVID-19 cases out of the total absenteeism cases varied throughout the study (being especially low at the start of the pandemic). However, by March 2020 (Wave-1), the Province of Alberta had the highest per capita rate of COVID-19 clinical testing performed of all Canadian Provinces (1371 per 100,000 vs 791 per 100,000) (Fletcher 2022). While the workforce studied represented the largest employer in the city, this prediction model may not be directly generalized to other workplaces and policies may differ across different institutions. Furthermore, within the workforce of this study, we could not distinguish the role individual affected workers fulfilled, and this may be an important future aspect to study as the risks of COVID-19 acquisition is heavily influenced by occupation (Shah et al. 2022).

## 5. CONCLUSIONS

These findings highlight the potential of WBS as a novel and objective approach that can monitor and predict workforce absenteeism related to COVID-19 infections with a one-week lead time. This model could be used to optimize workforce distribution to ensure efficiency and cost-effectiveness. WBS is an objective, inclusive, independent and cost-effective tool which can be leveraged to benefit society well beyond its currently focused healthcare-associated applications.

## Data Availability

All data used for this work is available through https://covid-tracker.chi-csm.ca or within the manuscript.

## AUTHOR CONTRIBUTIONS

**Nicole Acosta:** Formal analysis, Investigation, Methodology, Validation, Data Curation, Visualization, Writing - Original Draft, Writing - Review & Editing. **Xiaotian Dai**; Methodology, Formal analysis, Investigation, Writing - Review & Editing. **María A. Bautista**: Methodology, Validation, Investigation. **Barbara J. Waddell**; Methodology, Validation, Investigation. **Jangwoo Lee**; Investigation. **Kristine Du**; Investigation. **Janine McCalder**. Investigation. **Puja Pradhan**; Investigation. **Chloe Papparis**; Investigation. **Xuewen Lu**; Formal analysis. **Thierry Chekouo**; Formal analysis. **Alexander Krusina**; Investigation, Writing - Review & Editing. **Danielle Southern**; Investigation, Writing - Review & Editing. **Tyler Williamson**; Investigation, Writing - Review & Editing. **Rhonda G Clark**; Funding Acquisition, Project Administration. **Raymond Patterson**; Writing - Review & Editing. **Paul Westlund**; Writing - Review & Editing. **Jon Meddings**; Project administration, Writing - Review & Editing. **Norma J. Ruecker;** Project administration, Funding acquisition, Investigation, Writing - Review & Editing. **Christopher Lammiman**; Conceptualization, Investigation, Data Curation, Writing - Review & Editing. **Coby Duerr**; Conceptualization, Investigation, Data Curation, Writing - Review & Editing. **Gopal Achari:** Writing - Review & Editing. **Steve E. Hrudey**; Funding Acquisition, Formal analysis, Writing - Review & Editing. **Bonita E. Lee**; Funding Acquisition, Formal analysis, Writing - Review & Editing. **Xiaoli Pang**; Funding Acquisition, Formal analysis, Writing - Review & Editing. **Kevin Frankowski:** Conceptualization, Formal analysis, Writing - Review & Editing. **Casey RJ Hubert:** Conceptualization, Formal analysis, Funding acquisition, Writing - Review & Editing. **Michael D. Parkins:** Conceptualization, Formal analysis, Funding acquisition, Supervision, Writing - Review & Editing.

## ACKNOWLEDGMENTS

The investigators are grateful to the staff of the City of Calgary Water Services for their support in providing wastewater samples for assessment. We would also like to also thank the Calgary Emergency Management Agency (CEMA) team for collecting and sharing anonymous absenteeism case data. The authors are grateful for the efforts of the sample collection and preparation team including Alex Buchner Beaudet, Lawrence Man, Navid Sedaghat, Jennifer Van Doorn, Kevin Xiang, Leslie Chan, Laura Vivas, Kashtin Low and Andra Bacanu. The team would also like to acknowledge the efforts of Dr Jordan Hollman and Dr Jianwei Chen.

