## Supplemental for "Wastewater-based surveillance can be used to model COVID-19-associated workforce absenteeism"

**1. SUPPLEMENTARY TABLES**

**Table 1S.** City of Calgary’s employee absenteeism cases that were classified as a function of COVID-19; unrelated, COVID-19-related and COVID-19 confirmed per week.

| Week number | Date (week starting) | Unrelated | COVID-19 related cases | Confirmed COVID-19 cases |
| --- | --- | --- | --- | --- |
| 27 | 29/06/2020 | 35 | 32 | 0 |
| 28 | 06/07/2020 | 40 | 35 | 1 |
| 29 | 13/07/2020 | 53 | 26 | 0 |
| 30 | 20/07/2020 | 32 | 41 | 2 |
| 31 | 27/07/2020 | 53 | 42 | 0 |
| 32 | 03/08/2020 | 27 | 31 | 3 |
| 33 | 10/08/2020 | 43 | 43 | 0 |
| 34 | 17/08/2020 | 63 | 30 | 2 |
| 35 | 24/08/2020 | 49 | 41 | 0 |
| 36 | 31/08/2020 | 46 | 47 | 2 |
| 37 | 07/09/2020 | 41 | 37 | 0 |
| 38 | 14/09/2020 | 52 | 46 | 1 |
| 39 | 21/09/2020 | 59 | 73 | 3 |
| 40 | 28/09/2020 | 59 | 98 | 2 |
| 41 | 05/10/2020 | 53 | 63 | 3 |
| 42 | 13/10/2020 | 39 | 60 | 2 |
| 43 | 20/10/2020 | 59 | 91 | 3 |
| 44 | 26/10/2020 | 45 | 96 | 5 |
| 45 | 02/11/2020 | 50 | 94 | 8 |
| 46 | 09/11/2020 | 39 | 139 | 3 |
| 47 | 16/11/2020 | 53 | 118 | 15 |
| 48 | 23/11/2020 | 49 | 120 | 17 |
| 49 | 30/11/2020 | 55 | 141 | 23 |
| 50 | 07/12/2020 | 36 | 150 | 32 |
| 51 | 14/12/2020 | 40 | 118 | 29 |
| 52 | 21/12/2020 | 30 | 65 | 8 |
| 53 | 28/12/2020 | 12 | 51 | 12 |
| 2 | 04/01/2021 | 45 | 57 | 15 |
| 3 | 11/01/2021 | 58 | 59 | 18 |
| 4 | 18/01/2021 | 51 | 61 | 11 |
| 5 | 25/01/2021 | 44 | 64 | 9 |
| 6 | 01/02/2021 | 56 | 53 | 4 |
| 7 | 08/02/2021 | 52 | 42 | 6 |
| 8 | 15/02/2021 | 44 | 27 | 3 |
| 9 | 22/02/2021 | 55 | 50 | 7 |
| 10 | 01/03/2021 | 62 | 45 | 5 |
| 11 | 08/03/2021 | 58 | 46 | 7 |
| 12 | 15/03/2021 | 50 | 52 | 10 |
| 13 | 22/03/2021 | 44 | 71 | 18 |
| 14 | 29/03/2021 | 44 | 65 | 8 |
| 15 | 05/04/2021 | 35 | 137 | 25 |
| 16 | 12/04/2021 | 38 | 109 | 29 |
| 17 | 19/04/2021 | 48 | 119 | 32 |
| 18 | 26/04/2021 | 49 | 127 | 41 |
| 19 | 03/05/2021 | 50 | 200 | 48 |
| 20 | 10/05/2021 | 36 | 186 | 45 |
| 21 | 17/05/2021 | 48 | 120 | 23 |
| 22 | 24/05/2021 | 43 | 54 | 12 |
| 23 | 31/05/2021 | 42 | 24 | 2 |
| 24 | 07/06/2021 | 49 | 28 | 3 |
| 25 | 14/06/2021 | 44 | 24 | 7 |
| 26 | 21/06/2021 | 44 | 21 | 2 |
| 27 | 28/06/2021 | 46 | 13 | 0 |
| 28 | 05/07/2021 | 49 | 24 | 1 |
| 29 | 12/07/2021 | 42 | 16 | 0 |
| 30 | 19/07/2021 | 34 | 45 | 5 |
| 31 | 26/07/2021 | 48 | 27 | 3 |
| 32 | 02/08/2021 | 36 | 21 | 5 |
| 33 | 09/08/2021 | 50 | 30 | 5 |
| 34 | 16/08/2021 | 43 | 29 | 7 |
| 35 | 23/08/2021 | 37 | 22 | 4 |
| 36 | 30/08/2021 | 43 | 32 | 4 |
| 37 | 06/09/2021 | 57 | 80 | 13 |
| 38 | 13/09/2021 | 56 | 92 | 22 |
| 39 | 20/09/2021 | 76 | 76 | 17 |
| 40 | 27/09/2021 | 57 | 79 | 16 |
| 41 | 04/10/2021 | 51 | 66 | 18 |
| 42 | 11/10/2021 | 45 | 54 | 5 |
| 43 | 18/10/2021 | 67 | 48 | 5 |
| 44 | 25/10/2021 | 66 | 65 | 10 |
| 45 | 01/11/2021 | 79 | 44 | 10 |
| 46 | 08/11/2021 | 66 | 41 | 5 |
| 47 | 15/11/2021 | 64 | 44 | 4 |
| 48 | 22/11/2021 | 51 | 47 | 9 |
| 49 | 29/11/2021 | 68 | 53 | 8 |
| 50 | 06/12/2021 | 66 | 42 | 4 |
| 51 | 13/12/2021 | 38 | 44 | 7 |
| 52 | 20/12/2021 | 50 | 91 | 34 |
| 53 | 27/12/2021 | 25 | 231 | 167 |
| 2 | 03/01/2022 | 33 | 247 | 169 |
| 3 | 10/01/2022 | 44 | 222 | 125 |
| 4 | 17/01/2022 | 26 | 143 | 99 |
| 5 | 24/01/2022 | 28 | 139 | 102 |
| 6 | 31/01/2022 | 69 | 189 | 142 |
| 7 | 07/02/2022 | 60 | 156 | 122 |
| 8 | 14/02/2022 | 60 | 46 | 36 |
| 9 | 21/02/2022 | 49 | 28 | 23 |
| 10 | 28/02/2022 | 33 | 37 | 30 |
| 11 | 07/03/2022 | 53 | 19 | 14 |
| 12 | 14/03/2022 | 60 | 19 | 18 |
| 13 | 21/03/2022 | 48 | 15 | 13 |
| 14 | 28/03/2022 | 63 | 54 | 47 |

**2 SUPPLEMENTARY FIGURES**

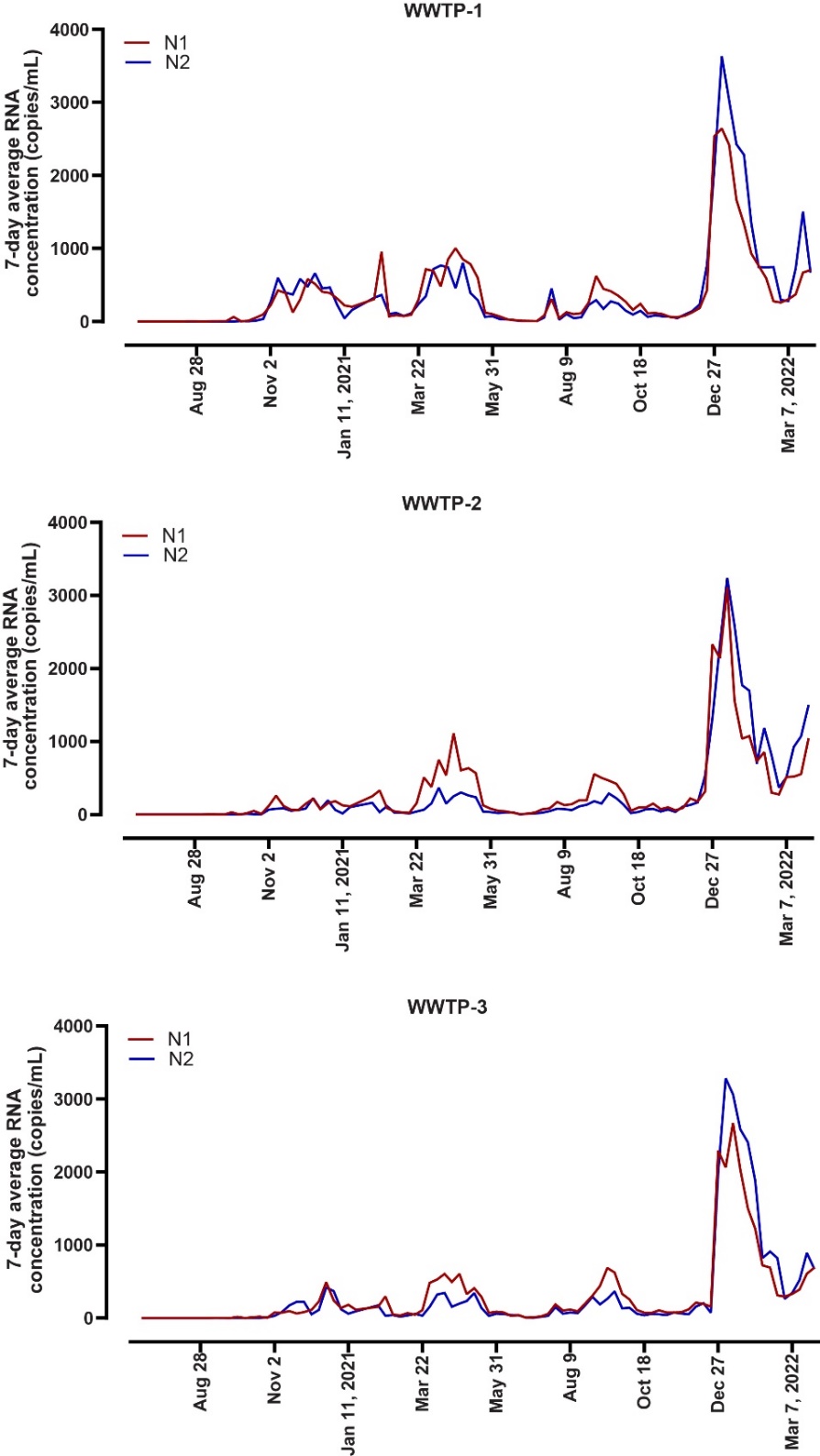

**Figure 1S. SARS-CoV-2 RNA concentration for Calgary’s three wastewater treatment plants (WWTPs).** Weekly average SARS-CoV-2 RNA signal (N1 (red line) and N2 (blue line)) from Calgary's three WWTPs: (A) WWTP-1, (B) WWTP-2 and (C) WWTP-3. Data were collected from June 29, 2020 to March 28, 2022 from all three WWTPs.

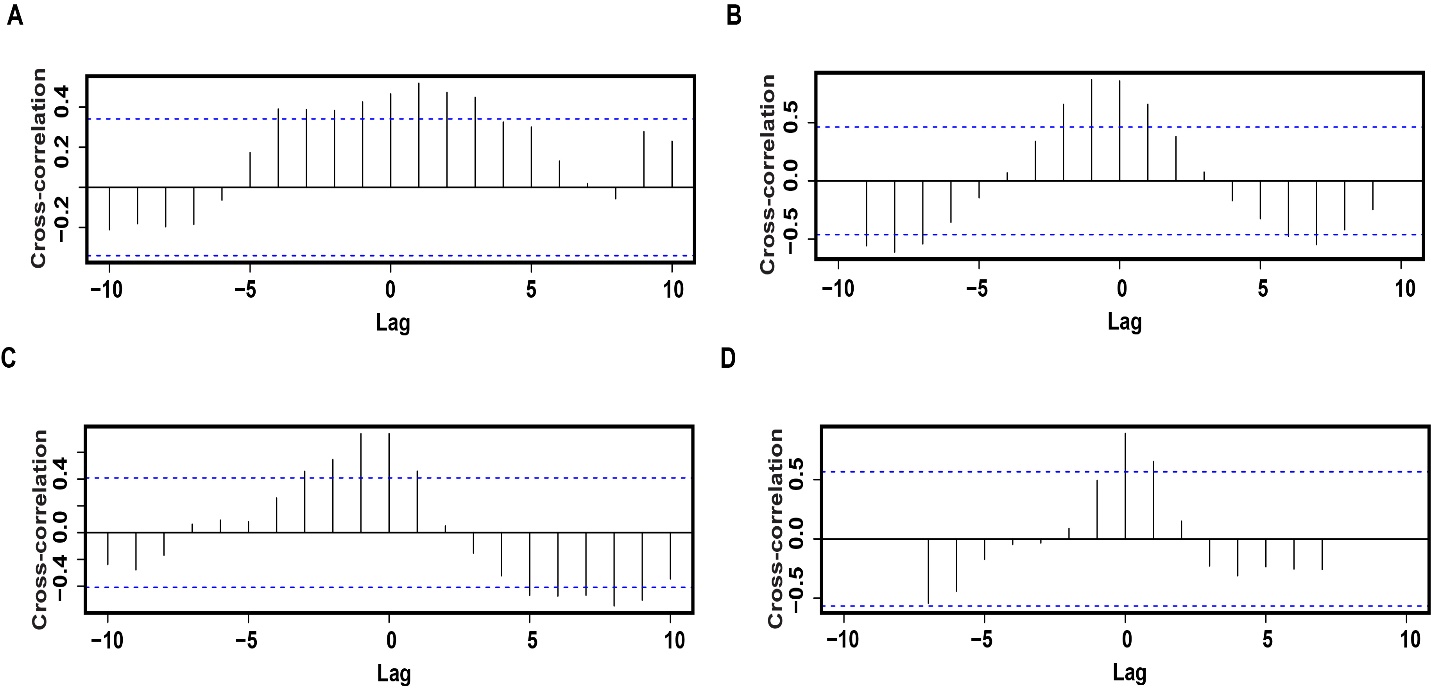

**Figure 2S. Cross correlation analysis for SARS-CoV-2 RNA in wastewater and municipal COVID-19-realted absenteeism**. RNA concentration measured with the N1 target and COVID-19-related absences during the Wave-2 (A), Wave-3 (B), Wave-4 (C) and Wave-5 (D). Blue doted lines denote the limits that determine the significance of the autocorrelation coefficients (95% confidence level).

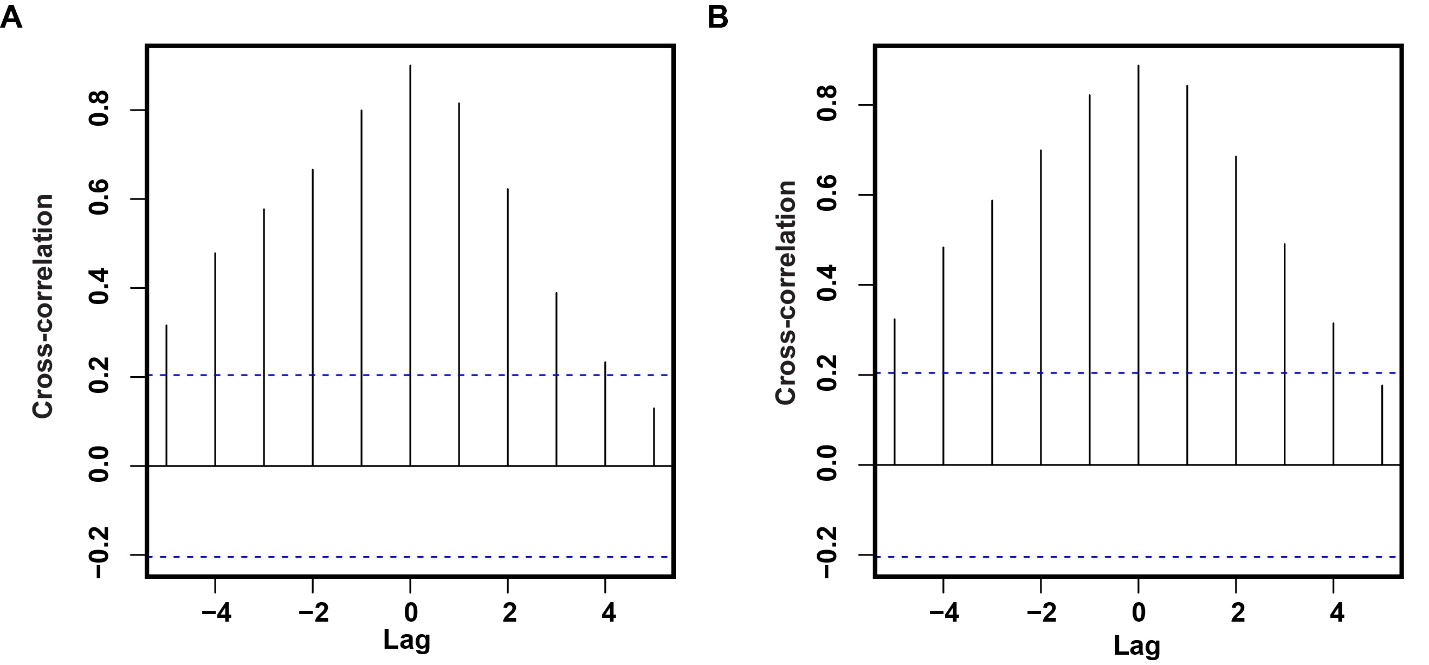

**Figure 3S. Cross correlation analysis for SARS-CoV-2 RNA in wastewater and municipal COVID-19-confirmed absenteeism**. RNA concentration measured either by the N1(A) or N2 (B) targets and COVID-19-confirmed absences. Blue doted lines denote the limits that determine the significance of the autocorrelation coefficients (95% confidence level).

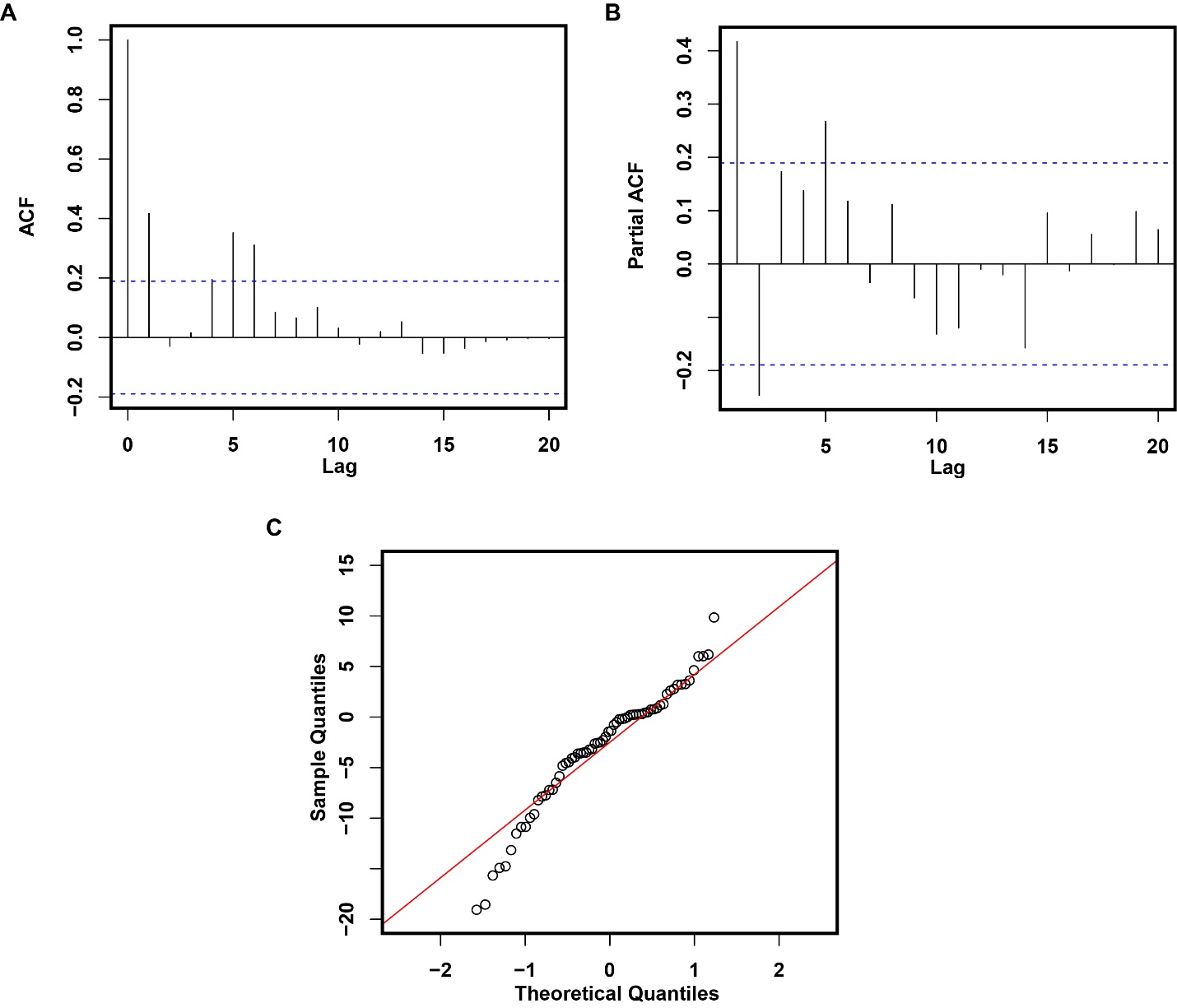

**Figure 4S. Residual diagnostics plots from the prediction model.** Autocorrelation (ACF) (A), Partial-correlation (PACF) (B) and normality (Quantile-quantile) plots from the residuals of the prediction model. Blue doted lines denote the limits that determine the significance of the autocorrelation or partial autocorrelation coefficients (95% confidence level).

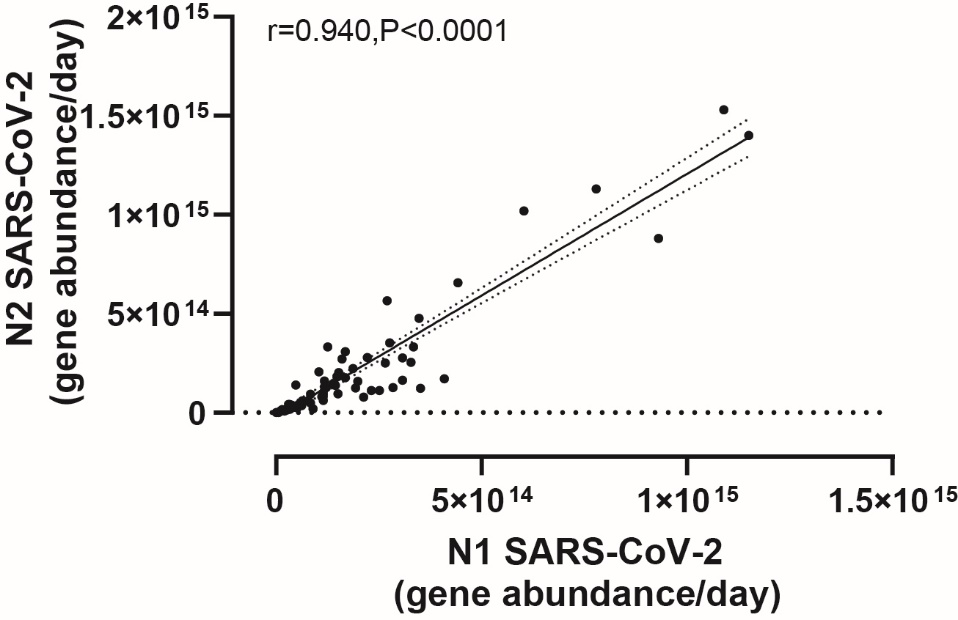

**Figure 5S. Comparison of SARS-CoV-2 RNA concentration in wastewater measured by the N1 and N2 targets.** Scatter plot of the weekly average of the gene abundance in wastewater measured with the N1 and N2 targets. Spearman correlation and 95% prediction intervals (dashed line) on the linear regressions (solid line) are shown in the figure.
